# LIVED EXPERIENCES OF MOTHERS WITH PERIPARTUM CARDIOMYOPATHY

**DOI:** 10.1101/2024.02.16.24302928

**Authors:** Doreen Macherera Mukona, Barbra Tsiko, Mathilda Zvinavashe

**Author notes:** Corresponding author, Doreen Macherera Mukona, Fatima College of Health Sciences, United Arab Emirates.

## Abstract

Peripartum cardiomyopathy is known to occur more commonly in African women or those of African descent, with potentially devastating consequences. The purpose of this study was to explore the lived experiences of women with post-partum cardiomyopathy. A phenomenological approach was used. The study was conducted with 6 women with a confirmed diagnosis of peri-partum cardiomyopathy who were registered with the Parirenyatwa Group of Hospitals Cardiac clinic. Excluded from the study, due to compromised autonomy, were the mentally ill, critically ill, and the institutionalized. Participants were selected as they came for routine management. In-depth interviews, following semi structured questionnaires, were conducted. All participants gave informed consent, and the study was conducted according to the requirements of the Declaration of Helsinki. Interviews were held in a private room and filled in questionnaires and detailed notes were anonymized and kept in a lockable cupboard to which the researchers had sole access. Manual thematic analysis was used to analyze the data and it was presented in themes, subthemes, and codes. The stages of thematic analysis followed were data organization, familiarization, transcription, coding, identifying themes, indexing, reviewing themes, displaying, and reporting. Trustworthiness was ensured by observing credibility, dependability, confirmability, and transferability. Three major themes namely: experience on diagnosis, psychosocial tension and supporting factors were identified. Women diagnosed with peripartum cardiomyopathy experience physical, psychological, and emotional stress. Psychosocial support is very vital in the management of peripartum cardiomyopathy.

## Introduction

Peripartum cardiomyopathy (PPCM) is the most common cardiomyopathy associated with pregnancy. Often heart failure can occur between the last month of pregnancy and the fifth month after childbirth. The etiology is undeterminable, with no previous evidence of heart failure.(Patel et al., 2016a) It is characterized by cardiac failure secondary to left ventricular malfunction towards the end of the third trimester or five months after giving birth. The left ventricle may not be dilated, but the ejection fraction is significantly reduced to as low as 45%.(Agarwal et al., 2019) Normal values of ejection fraction range from 55-65 percent. This leads to reduced perfusion of all vital organs including the lungs, liver, and other systems of the body. The major signs of peripartum cardiomyopathy include heart failure, shortness of breath and generalized edema. In pregnant women diagnosis is often skipped or delayed as symptoms are close to those of regular pregnancy hemodynamic changes. Other complications include hypoxia thromboembolic, progressive heart failure, arrhythmias, fetal distress due to maternal hypoxia and placental hypo perfusion and maternal hypovolemia.(Arany and Elkayam, 2016)

Due to lack of population based studies, the true incidence or prevalence of PPCM in Africa is unknown.(Karaye et al., 2018) However, it is more common in black women.(Arany and Elkayam, 2016) Hospital-based studies conducted in Africa have yielded values such as 1 in 1000 live births in South Africa, 1 in 100 deliveries in Sokoto, Nigeria, and 1 in 3,800 in Burkina Faso.(Desai et al., 1995, Isezuo and Abubakar, 2007, Yaméogo et al., 2018) The occurrence of postpartum cardiomyopathy is about 1 in 2 500-4 000 births in the United States America. The prevalence is estimated to be 1 case per 1 000 live births in South Africa, and 1 case per 350-400 live births according to international statistics. Though PPCM can occur at any age, about half of cases occur in older women over 30 years of age. A case control study found that black women had a 15.7 fold higher risk of peripartum cardiomyopathy compared to non-black women.(Arany, 2020) In Sub-Saharan Africa, heart diseases, including PPCM, have reached epidemic proportions and is a significant contributor to morbidity and mortality. These conditions disproportionately affect young women particularly during pregnancy, and they have a worse prognosis than other groups of patients. This has far reaching effects for the patient, children and the whole family unit.(Sliwa et al., 2010)

Risk factors for PPCM include increased age, gravidity or parity, African origin, preeclampsia, use of tocolytics, twin pregnancy, obesity, low socioeconomic status, customary birth practices, malnutrition, and selenium deficiency.(Karaye et al., 2016) Understanding “what it is like” from the point of view of the sufferer has been recognized as a fundamental starting point for caring. It is necessary not only to provide empirical awareness of the state of a patient, but also to consider the subjective life experience and sense of illness from the viewpoint of the sick individual.(Fett, 2013)

Though PPCM is known to occur more frequently in African women or those of African descent, there is very little reported data on their lived experiences. Moreover, the effects of PPCM are potentially overwhelming. No studies have been conducted in Zimbabwe to explore the lived experiences of women with PPCM. The experiences are likely to be worsened by poorly resourced public hospitals that render it suboptimal to diagnose and treat these patients.(Munyandu et al., 2017) A quick records review conducted by the researcher at the study site, revealed a 12-15 % prevalence of PPCM among patients seen in Outpatients Department Cardiac. A registry data base of patients with PPCM kept at the site, had 200 patients at the time this study was conducted.

It is very important for midwives to understand the lived experiences of women with peripartum cardiomyopathy, because it results in significant morbidity and mortality.(Main et al., 2015) Health care laws and guidelines in Zimbabwe emphasize the importance of taking the expertise, perspectives and expectations of the patient into account and discerning the particular needs of the client in their own care.(Munyandu et al., 2017, Gambahaya et al., 2022) Healthcare is needed to alleviate pain, increase the standard of care in relation to pregnancy and childbirth, relieve distress and enhance the quality of treatment. As such, health care professionals/midwives must consciously listen to the narratives of patients to provide individualized and integrated quality care. The purpose of this study, therefore, was to explore the lived experiences of women diagnosed with peripartum cardiomyopathy at Parirenyatwa Group of Hospital in Zimbabwe.

## Methodology

The purpose of this study was to explore the lived experiences of women with peri-partum cardiomyopathy. A phenomenological approach was used. The study was conducted with 6 women with a confirmed diagnosis of post-partum cardiomyopathy. These women were registered with the Parirenyatwa Group of Hospitals Cardiac clinic. Excluded from the study, due to compromised autonomy, were the mentally ill, critically ill, and the institutionalized. Participants were selected as they came for routine treatment and management at the site. Data were collected through in-depth interviews following semi structured questionnaires from the 23^rd^ of July 2020 to the 7^th^ of August in the same year. All participants gave informed consent by signing the consent form, and the study was conducted according to the requirements of the Declaration of Helsinki. A witness to the consent process also signed. Interviews were held in a private room and filled in questionnaires and detailed notes were anonymized. They were then kept in a lockable cupboard to which the researchers had sole access. Manual thematic analysis, as recommended by Braun and Clarke was used to analyze the data and it was presented in themes, subthemes and codes. (Clarke and Braun, 2017) The stages of thematic analysis followed were data organization, familiarization, transcription, coding, identifying themes, indexing, reviewing themes, displaying and reporting. Trustworthiness was ensured by observing credibility, dependability, confirmability, and transferability.

### Findings

Table 1 presents the demographic characteristics of the participants. The gravidity, parity and total number of living children ranged from 1-4.

**Table 1:** Demographic characteristic of participants (n=6)

| Participants (P) | 1 | 2 | 3 | 4 | 5 | 6 |
| --- | --- | --- | --- | --- | --- | --- |
| Age range in years | 30-34 | 35-39 | 20-24 | 25-29 | 30-34 | 40-44 |
| Marital status | Married | Divorced | Divorced | Married | Married | Married |
| Level of education | Primary | Secondary | Secondary | Secondary | Tertiary | Secondary |
| Employment | Formally Employed | Unemployed | Self employed | Self employed | Formally employed | Formally employed |
| Monthly income in Zimbabwean dollars | \$2000.00 to \$2900.00 | 0-\$1000.00 from donations | 0-\$1000.00 | 0-\$1000.00 | \$3000.00 | Above \$3000.00 |
| Area of residence | Urban | Urban | Urban | Urban | Urban | Rural |
| Religion | Christian | Christian | Christian | Christian | Christian | Christian |

Table 2 presents codes and subthemes relating to theme 1 (experience with the initial diagnosis).

**Table 2:** Theme 1 Experience with the initial diagnosis.

| Themes | Sub themes | Codes |
| --- | --- | --- |
| Experience with the initial diagnosis | Initial symptoms | Cough, shortness of breath, swollen face and legs, fatigue, inability to walk, chest pain, general body swelling, dyspnea on exertion, inability to lift or carry the baby.<br>Inability to carry out household chores and caring for the babies.<br>Symptom onset ranging from two days to 3 months.<br>Intensive Care Unit (ICU) admission. |
|  | Symptom dismissal | Dismissal of symptoms as normal occurrence after birth<br>Symptom dismissal as attention seeking tendencies |
|  | Inaccurate diagnosis | Wrong diagnosis of high blood pressure<br>Wrong diagnosis of asthma<br>Wrong diagnosis of pulmonary embolism<br>Wrong diagnosis of anemia |

Table 3 presents the codes and subthemes relating to theme 2 (Psychosocial tension)

**Table 3:** Theme 2 Psychosocial tension.

| Themes | Sub themes | Codes |
| --- | --- | --- |
| Psychosocial tension | Initial reaction | Overwhelming shock of receiving the diagnosis<br>Difficulty accepting the diagnosis<br>Devastating nature of the diagnosis<br>Hurt<br>Worry about perceived impending death and leaving a |
|  |  | newborn behind<br>Denial of diagnosis<br>Depression from separation from baby due to ICU admission and failure to breastfeed<br>Acceptance after explanation by the doctor and counselors |
|  | Impact on childbearing and family planning | Certainty of high-risk pregnancies with decision to have more children<br>Limited options for family planning<br>Forced option for permanent methods of family planning<br>Gloom over shattered intentions to have more children<br>Option for long term methods of family planning<br>Abnormal fear of falling pregnant. |
|  | Strained relationships | Job loss due to prolonged absence from work and disability<br>Financial difficulties<br>Lack of understanding of condition by some relatives<br>The beggar status |

Table 4 presents the codes and subthemes relating to theme 3 (Supporting factors)

**Table 4:**
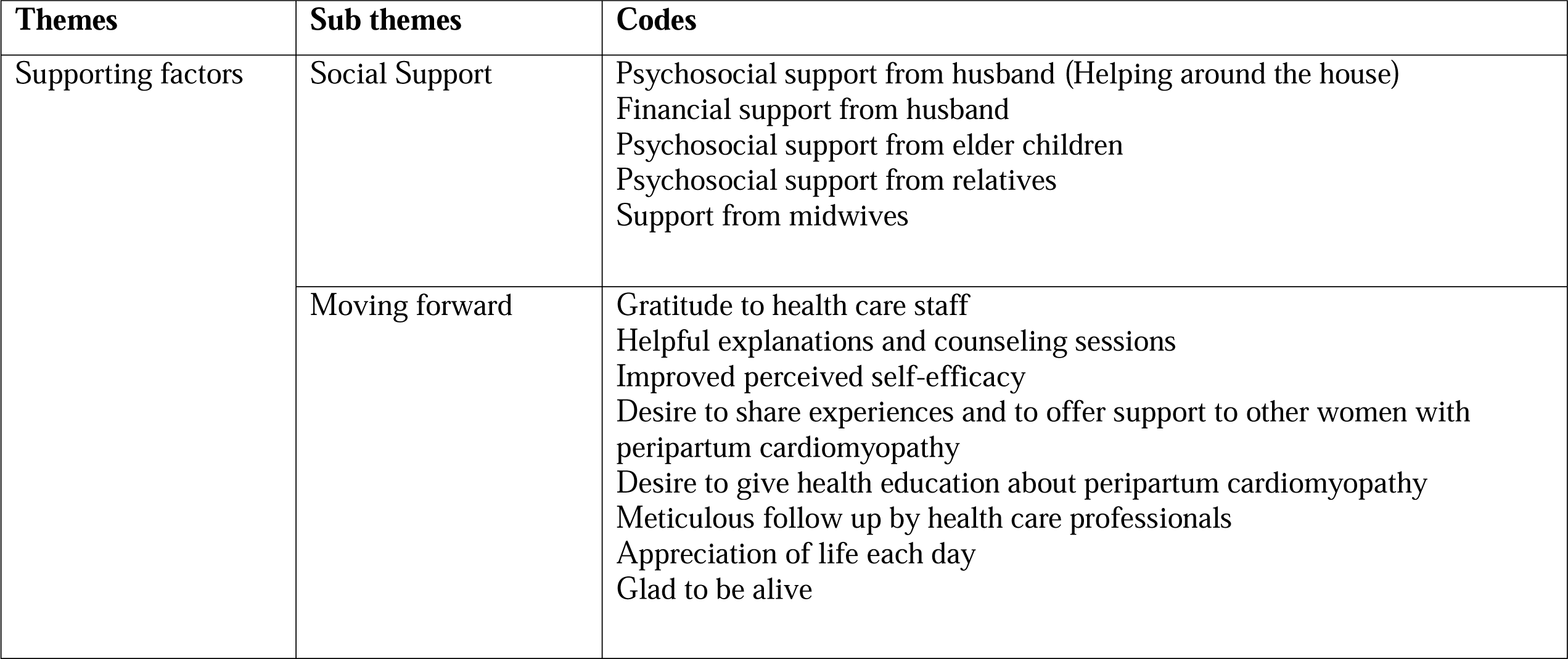
Theme 3 Supporting factors.

### Experience with the initial diagnosis

The first theme identified during was our study was the experience with the initial diagnosis. All participants narrated how they experienced symptoms such as coughing and shortness of breath. The sub-themes building up to this theme, therefore, were initial symptoms, symptom dismissal and inaccurate diagnosis.

#### Initial symptoms

Participants narrated several symptoms they experienced prior to their diagnosis. These included cough, shortness of breath, chest pain, intense anxiety, over-swelling of the body, and inability to perform daily tasks. Some participants had this to say:

> *‘I started coughing, experiencing shortness of breath, swollen face and legs, inability to carry out household chores and caring for my babies and this started three months after delivery of my twins. I thought I had high blood pressure.’* P1

> ‘*I started having shortness of breath, general body swelling, fatigue and unable to walk to nursery to feed my baby a week after delivery of a preterm baby by caesarian section because of pregnancy induced hypertension.’*P2

> ‘*Three months after my second daughter was born, I started coughing, difficulty in breathing when going up the stairs, and I felt heavy in the chest when carrying my baby. I thought I had developed asthma.’*P4

#### Symptom dismissal

Some participants narrated how their symptoms were dismissed by either themselves or by health staff as normal occurrences after giving birth. This is captured in the following excerpts:

> *‘My legs were swollen, and it was difficult to walk. I thought it was high blood pressure from the twin pregnancy.’*P1

> ‘*Three months after my second daughter was born, I started coughing, difficulty in breathing when going up the stairs, and I felt heavy in the chest when carrying my baby. I thought I had asthma since my mother is asthmatic.’* P4

> *’I began to experience extreme shortness of breath two days after my son’s birth, so it was hard f or me to get out of bed. I mentioned this to a nurse who said you have pulmonary embolism, of course, the doctor said.’* P5

#### Inaccurate diagnosis

Some participants reported how they got wrong diagnoses before the correct one of peri-partum cardiomyopathy was made. One participant had this to say:

> *‘I went back to the rural clinic where I had delivered two weeks later experiencing shortness of breath, swollen face and legs, the nurse said ‘You have anemia because you have delivered and have lost blood, you need to take iron tablets, which she prescribed for me. I did not get better and went to see a doctor in Bindura who referred me to a cardiologist, who made the accurate diagnosis later.’* P6

### Psychosocial tension

The second theme identified in this study was psychosocial tension after the diagnosis of peripartum cardiomyopathy. Participants narrated feelings of loss, anger, hurt and depression. Some reported being nervous and scared. Psychosocial tension was also brought about by the burden of rethinking fertility intensions and strained social relationships. The subthemes identified were the initial reaction, impact on childbearing and family planning and strained relationships. Some participants narrated the following:

#### The initial reaction

Experience with the initial diagnosis of PPCM was mostly met with shock. Some participants reported feeling hopeless. Some felt scared of the possibility of death and leaving a new-born baby. Participants had this to say:

> *’I was really worried because I felt I might die and leave my first child behind.* ‘*I was very much worried because I thought I was going to die and leave my first child. My husband divorced me because I was very sick. I recovered and had stopped taking drugs. I fell pregnant by mistake from another man and delivered a second baby. I started coughing, having difficulty in breathing and swollen legs. I was put back on drugs for the heart for the second time. I thank God today after one year follow up echocardiography doctor said I have recovered and stopped the medication.’* P3

> *‘I am very depressed from being separated from my baby while being admitted in ICU and not breast feeding my son.’* P5

#### Impact on childbearing and family planning

Some participants narrated difficulties in accepting the sudden limited family planning options on top of the diagnosis of peripartum cardiomyopathy. To some the diagnosis meant that they could not have either a normal pregnancy again, or they could not have any more children. This is captured in the following excerpts:

> *‘The doctor explained to me and my husband that my heart was so weak, and it is risky to fall pregnant again because with some women the condition relapse and some women the peripartum cardiomyopathy improves. I have opted for sterilization as a method of family planning since I now have four children.’* P1

> *‘I’m very sad that my husband and I are not going to have any more kids. This is totally damaging*. *We have opted for an intra-uterine device as a method of family planning.’*P5

> P 6 said *‘I was scared of falling pregnant again. My husband and I agreed to have my tubes tied. Those three children are enough. We followed the doctor’s advice though he said my heart is improving.’*

#### Strained relationships

Some participants experienced strain in their relationships as family members could not understand the sudden incapacitation after a normal birth. Some lost their jobs due to extended periods of absenteeism from work. This is reflected in the following excerpts:

> *‘It is very difficult for me because I lost my job because of disability. The little money which I get from my ex-husband is not enough for food and drugs. My relatives always say “how can you buy drugs when there is no food in the house.” At times I default treatment because I will not be having enough money to buy drugs because they are very expensive and some need to be purchased in United States dollars. Some if the investigations are expensive especially echocardiography. Most of the time I depend on donations from the church.’* P2

> *‘It was difficult for me at first because mother-in-law could be angry with me when my husband helped with preparing meals for the family but later when my husband explained my condition to her, she became so supportive. Medications are very expensive, and we sometimes travel to Harare to buy them. If we can get them locally it could save us.’* P6

### Supporting factors

The last theme identified was “supporting factors.” The sub-themes were social support and moving forward.

### Social support

Some participants expressed gratitude to family and health staff for the care and support provided before and after the diagnosis. One participant had this to say:

> *‘My husband is very supportive. He always buys the medications for me although they are very expensive. He does most of the cooking laundry and cleaning of the house. My elder children also assist with caring for the babies and other household chores and our niece has moved in to stay with us so that she can also help.’* P1

### Moving forward

Participants in this study expressed gratitude for the health education and support received by spouses, family members, and health care professionals. This gave them hope and enhanced their self-efficacy to manage their conditions as well as supporting other women with peripartum cardiomyopathy. Some were even thinking of forming support groups and using social media for wider coverage. Some participants had this to say:

> *’I am grateful to all the health care professionals who have taken care of me during my hospital stay.’ The physician explained everything to me and I do not hesitate to move forward in this condition. I am prepared to share with other women on WhatsApp groups my experience with peripartum cardiomyopathy.’*P3

> *‘I am grateful for the education I receive and the follow up care from our doctor, she takes her time to explain everything. This gives me courage to move forward.’* P4

> *’I am grateful that I have lived to see the sixth birthday of my son.*

> ‘*I am thankful to be alive. I am willing to form a support group with other mothers so that we can share our experience and offer encouragement to move forward.’* P6

## Discussion

This was a phenomenological study conducted with 6 women with peripartum cardiomyopathy at a central hospital in Harare, Zimbabwe. Ages of the participants ranged from 24 years to 40 years. Peripartum cardiomyopathy is common in women above 30 years of age,(Sliwa et al., 2014) though it can occur at any age. Sociodemographic factors such as age, marital status, parity, and religion can significantly alter a person’s response to illness and, subsequently, the prognosis. Other risk factors for peripartum cardiomyopathy may include lower economic group with poor nutritional status, multiparity and history of peripartum cardiomyopathy. Other studies have reported lack of formal education, unemployment, underweight and history of pre-eclampsia as independent risk factors for peripartum cardiomyopathy.(Karaye et al., 2020a) Women who are obese, African, or have a history of high blood pressure, multiple pregnancies, and smoking are at elevated risk of developing peripartum cardiomyopathy.(Dekker et al., 2016) The disease is generally more prevalent in Asia and Africa.(Mubarik et al., 2018)

### Initial symptoms

Mothers diagnosed with PPCM experience distinct memories of their initial symptoms and whether they are taken seriously by themselves, their family/friends, midwives and other health professionals, as symptomatology is easily mistaken with “natural late pregnancy or postpartum pain.”(Fett, 2013) Peripartum cardiomyopathy is often associated with severe heart failure occurring towards the end of pregnancy or in the months following birth with debilitating, exhausting and frightening symptoms requiring person-centered care.(Patel et al., 2016b, Patel et al., 2016a) Capturing the PPCM diagnosis experience of mothers is very important for midwives, since peripartum cardiomyopathy is associated with severe morbidity, including ventricular dysrhythmias, thromboembolism and recurrent.(Fett, 2013) It is often associated with feelings of devastation, extreme fear, doom, hysteria, freaking out and emotional torture.(Dekker et al., 2016) In our study, some women had health care training backgrounds, but the shock of hearing the diagnosis and prognosis was daunting even for them. Findings of a study conducted in Sweden also revealed exacerbated suffering related to failing health symptoms, not being cared for and not feeling healthy. It is very vital to adopt a bio-psycho-social, person-centered approach in the care of women with PPCM.(Ekman et al., 2011) Participants in this study cited complete exclusion of PPCM among differential diagnoses even when mothers presented with complaints of severe orthopnea, chest pain, swollen abdomen, lower limb edema and unable to do any job. This resulted in delayed diagnosis and treatment. Similar findings have also been reported by Hess et al. and Dekker and colleagues.(Hess and Weinland, 2012, Dekker et al., 2016) When explaining seeking care for peripartum cardiomyopathy symptoms, Hess et al. found that women used words such as “dismissed and brushed off.” PPCM was often mistaken for hypertension caused by pregnancy, anemia, post-delivery hypertension and post-caesarian pulmonary embolism, which led to the delay in treatment.(Hess and Weinland, 2012) Worse still, cardinal symptoms in PPCM such as shortness of breath, dizziness and edema can occur in multiple pregnancy or in normal pregnancies.(Ramaraj and Sorrell, 2009) In this study participants experienced initial misdiagnosis such as hypertension, anemia, and pulmonary embolism. Misdiagnosis of PPCM can have grave consequences. One study conducted in California reported 13 out of 29 maternal dilated cardiomyopathy deaths (45%) being directly due to misdiagnosis of PPCM and subsequent treatment delays.(Harper et al., 2012) Though the clinical course can range from mild symptoms of heart failure to severe forms with cardiogenic shock,(Sieweke et al., 2020) the condition can be quite manageable when diagnosed and treated early.(Pfeffer et al., 2020) Management aims at optimizing hemodynamic status with sodium and fluid restriction, light exercise and oral heart failure therapy.(Sliwa et al., 2014)

It is, therefore, important for midwives working with pregnant and postnatal mothers to be mindful of peripartum cardiomyopathy symptoms to minimize symptom dismissal and enhance good prognosis in mothers with peripartum cardiomyopathy. PPCM should always be included among the differential diagnoses in mothers with cardiac-related complaints.(Regitz-Zagrosek et al., 2011) Globally, according to experts and national guidelines, the classic signs of heart failure in a woman towards the end of pregnancy or in the first months post-partum are an indication for clinical examinations such as electrocardiogram (ECG), chest X-ray and echocardiogram.(Regitz-Zagrosek et al., 2011)

### Psychosocial tension

Generalized anxiety, cardiac anxiety, and quality-of-life concerns are rampant in patients with PPCM at all stages of recovery. These feelings fueled by the rarity, nature of the presentation, and acuity of the illness. (Koutrolou-Sotiropoulou et al., 2016) Besides, the affected women are in most cases in their prime years with a lot of responsibilities and plans to have more children and, suddenly, they get limited by a heart problem.(Koutrolou-Sotiropoulou et al., 2016) Psychosocial issues are real and need to be addressed in patients with PPCM. (Rosman et al., 2019) PPCM imposes both an emotional and a physical burden especially on young mothers, years after the diagnosis. Findings of an earlier study conducted online revealed that almost two thirds participants felt discouraged with only a quarter reporting being satisfied with the counseling from health-care providers.(Koutrolou-Sotiropoulou et al., 2016) There are also many unplanned economic repercussions that are related to peripartum cardiomyopathy. In this study some participants were not formally employed while one lost her job due to ill health. The financial problems are overwhelming and worrisome. Some participants defaulted treatment because they could not afford to buy medications. Becoming a beggar due to illness imposes a huge psychological burden on a previously self-reliant individual. One participant even reported that she was divorced by her spouse due to the illness. This is a huge psychological burden that can potentially invoke a host of negative emotional reactions that include hopelessness and depression.(Koutrolou-Sotiropoulou et al., 2016) A study on husbands of women with PPCM revealed that husbands, in some instances, are equally shattered by the diagnosis. They also experience shock at the sudden overwhelming news, distress due to the uncertainty of the outcome and the future, helplessness when they are required to be strong, and disappointment.(Patel et al., 2019)

Some participants in our study expressed fear of ever falling pregnant again. The fears are quite substantiated as data suggests that a relapse of PPCM happens in about one third of women who get pregnant.(Dekker et al., 2016) Some studies have reported major mental health disorders with PPCM. (Pfeffer et al., 2020)A study conducted earlier at the same site as our study reported a mortality rate of 11.6% over a year among women with peripartum cardiomyopathy(Munyandu et al., 2017) while another study conducted in Nigeria reported a mortality of 18.7%.(Karaye et al., 2020b) One participant, in our study, who unintentionally got pregnant, experienced a relapse of peripartum cardiomyopathy with the subsequent pregnancy. The possibility of peripartum cardiomyopathy is related to the current cardiac function of a mother and thus individualized.(Elkayam, 2014) It is very important to explain the pros and cons of different birth control methods, the potential for relapse and the value of preventing pregnancy while still maintaining a reduced ejection fraction. (Fett et al., 2015) In this study almost all participants opted for long term methods of family planning such as intrauterine device and tubal ligation to prevent pregnancy. Estrogen-progestin contraceptives may increase fluid retention, which may make HF worse. Early after diagnosis, and in women with chronic left ventricular dysfunction, combined oral contraceptives tend to increase the risk of thromboembolism.(Curtis et al., 2016, Tepper et al., 2019)

### Supporting factors

Women are very vulnerable after PPCM diagnosis and struggle to find psychological balance. However, the process of psychological adaptation after PPCM, remains largely unexplored. Professional psychological support should be included in the overall management plan rather than giving highest priority to physical symptoms alone.(de Wolff et al., 2018) Some participants in our study reported that they could only accept their condition after careful explanation and counseling regarding the diagnosis of PPCM. News about the diagnosis of peripartum cardiomyopathy needs to be provided in a sensible way. Accurate, evidence-based information on their mortality risk and prognosis for recovery is very important. This empowers the affected women to accept their condition. Knowledge of the disease and medical management might help in recovery.(Koutrolou-Sotiropoulou et al., 2016) Participants in this study considered the diagnosis of peripartum cardiomyopathy to be a devastating experience. However, they applauded physicians for giving them accurate information concerning their diagnosis and treatment management and offered psychological support on how to move forward. Their faith and religion helped them to accept their conditions. Some mothers were assisted by family members and some donations from the church to cover hospital bills and medications. Some coping mechanisms reported in other studies are physical activity, positive thinking, professional counseling, career changes, and furthering education.(de Wolff et al., 2018) In some studies, even partners of affected women expressed gratitude towards health care professionals their support.(Patel et al., 2019)

## Conclusion

Women with peripartum cardiomyopathy go through a range of experiences that go beyond physical illness. The physical illness itself might provoke other psychosocial issues such as financial problems, strained relationships, and the need to rethink fertility options. They also go through significant psychological tension, and they need psychosocial support to enhance their adjustment to the new diagnosis.

## Data Availability

Data available on reasonable request

